# Participant attitudes toward returning individual results from CADASIL research

**DOI:** 10.64898/2026.08.21.26361045

**Authors:** Deven K. Burks, Elizabeth Penziner, Lindsay R. Clark, Fred B. Ketchum, Kenneth D. Croes, Jane S. Paulsen, United States CADASIL Consortium

**Author notes:** Corresponding author: Deven K. Burks, 1685 Highland Avenue, Madison, WI, 53705, United States of America.

## Abstract

**INTRODUCTION:** Neurodegenerative research identifies biomarkers to confirm presence of disease and inform about risk for clinical symptoms. Expert guidance advises caution about disclosing individual research results (IRR), but participant interest remains high even when IRR may not inform individual prognosis. Existing studies of stakeholder attitudes emphasize Alzheimer’s disease (AD) biomarkers. We explore participant attitudes toward IRR from the United States CADASIL Consortium (USCC), an observational study of Cerebral Autosomal Dominant Arteriopathy with Subcortical Infarcts and Leukoencephalopathy (CADASIL), the most heritable form of vascular dementia.

**METHODS:** Since CADASIL research participant attitudes are unstudied and AD-focused guidelines for IRR may not generalize to populations with dominantly inherited conditions, we surveyed USCC participants using three 5-point Likert items and one open-ended question. Descriptive statistics were analyzed for Likert items. The distribution of responses to one item was directly compared to an AD participant survey. Open-ended responses underwent qualitative content analysis.

**RESULTS:** We received 152 responses. The highest-rated reason to return IRR was “learn about my disease and its predicted course”. The highest-rated IRR were imaging/MRI scans and cognitive testing. Hypothetical negative outcomes were rated as a little to somewhat concerning. USCC respondents rated reasons to return IRR higher than AD counterparts, with statistically significant differences for seven of eight items. In open-ended responses, the most frequent code was “IRR return will help improve my health and well-being”.

**DISCUSSION:** Most respondents expressed support for disclosure upon participant request. These findings could inform IRR guidance for CADASIL and other disorders and investigations of personal utility.

## 1. Introduction

Translational research identifies biomarkers that confirm presence of disease and inform about risk for clinical symptoms. In neurodegenerative research, markers may be available before clinical symptom onset: biofluid assays and positron emission tomography (PET) imaging of amyloid-beta and tau in Alzheimer’s disease (AD); biofluid assays of neurofilament light chain and MRI volumetrics in amyotrophic lateral sclerosis (ALS), Huntington’s disease (HD), and frontotemporal dementia (FTD); age of HD motor diagnosis and functional decline. Consequently, existing or new risk markers may offer substantial information.

Nevertheless, translational research has outpaced person-centered research about disclosing individual research results (IRR). In the United States (US), expert committees have described ethical, legal, and social implications of communicating IRR to study participants.(1–8) Generally, experts agree that IRR may be returned when valid, interpretable, and actionable. Since research findings need not meet Food and Drug Administration standards for biomarker qualification, findings are not typically considered validated by consensual, recommended criteria.(9) Recent testing guidelines emphasize AD, yet cross-disease applications are constrained by distinct symptoms, severity, course, prominence, or populations. The applicability of these guidelines to IRR from studies of dominantly inherited neurodegenerative disease is particularly uncertain, given the dearth of studies in these populations.(10) Consequently, findings and guidelines from sporadic AD may not generalize across neurodegenerative diseases, particularly for dominantly inherited conditions.

For some stakeholders, the importance of returning IRR is evident. Disclosure could bolster study recruitment and retention, increase trust in science, and ensure research success.(11–21) Such claims remain speculative, absent studies to measure these outcomes. Additionally, disagreement persists among other stakeholders to what extent IRR benefit participant well-being. More research into stakeholder attitudes is warranted beyond sporadic AD. We therefore surveyed participants in the US CADASIL Consortium (USCC), an observational study of Cerebral Autosomal Dominant Arteriopathy with Subcortical Infarcts and Leukoencephalopathy (CADASIL), to explore their attitudes toward IRR, which are previously unstudied.

The USCC does not currently return IRR given the lack of therapeutic interventions, qualified biomarkers, regulatory approval, and budgetary/staffing support. Nonetheless, many participants have requested IRR, with several participants leaving the study citing dissatisfaction with the policy. Participant interest may stem from the clinical symptoms characteristic of CADASIL. A variant in the *NOTCH3* gene provokes recurrent strokes, cognitive decline, and psychiatric disturbances as early as the third and fourth decades of life. The variant location in the *NOTCH3* gene affects symptom severity and disease-course, with 300+ known variants of CADASIL.(22, 23)

## 2. Methods

To study CADASIL from presymptomatic through early disease stages, the USCC has enrolled 560 at-risk persons for a disease-causing *NOTCH3* variant. To be eligible, interested persons must be 18 years or older and have a known or suspected family history of CADASIL, with another family member having a confirmed *NOTCH3* variant. While many participants know their *NOTCH3* gene-status at enrollment, persons with a family history may enroll without genetic documentation, receive genetic counseling through the study, and choose to learn their gene-status or remain blinded. The study excludes persons with comorbid neurological conditions making findings uninterpretable, active uncontrolled diseases requiring medical attention for stability, or severe functional impairment (a modified Rankin Scale score >3, indicating loss of unassisted walking).

To understand USCC participant attitudes toward disclosure, we designed a four-item survey. The first five-point Likert item asked participants to rate each of twelve reasons for disclosure from not-at-all to extremely important. Eight reasons were adapted from previous AD surveys.(24, 25) Four reasons were developed from USCC participant feedback. The second five-point Likert item asked participants to rate each of ten data types from not-at-all to extremely important to return, adapted from US AD Research Center (ADRC) surveys.(14, 26) The third five-point Likert item asked participants to rate how concerned they would be, from not-at -all to extremely, in each of four scenarios where prognostic information was hypothetically returned, adapted from another AD survey.(24) The fourth item was open-ended: why participants should or should not be able to receive their IRR. Participants could choose not to respond to any items; non-responses were treated as missing values. Participants were not compensated for survey completion.

The survey was approved by the USCC’s centralized institutional review board (tracking number: 20216179) and loaded in Qualtrics (Seattle, WA). It was emailed to all USCC participants, with a two-week reminder. It was available 7/14/2025– 9/17/2025 and received 152 total responses. No demographic or clinical characteristics were collected for this anonymous survey.

Likert items were analyzed quantitatively. Qualtrics Data & Analysis tools generated descriptive statistical findings. For exploratory analysis, we compared the distribution of responses to equivalent items between this survey and a survey of the Wisconsin ADRC and Wisconsin Registry for Alzheimer’s Prevention (WRAP), observational cohort studies recruiting cognitively unimpaired individuals at risk for AD. The ADRC/WRAP survey respondents were aged 64.8 years (mean), 76% female, 56% White and 44% Black/African-American, and 58% bachelors-level education or higher.(24) Because survey responses were skewed, an independent samples Mann-Whitney U Test was conducted in IBM SPSS Statistics (v.30.0) (Armonk, NY) to compare importance ratings.

Two raters completed qualitative content analysis of open-ended responses (n=77) using spreadsheets. Insofar as stakeholder attitudes toward biomarker testing have previously been studied, content analysis took a hybrid approach.(27) Using a template approach,(28) a preliminary deductive codebook was developed from themes reported by comprehensive reviews of biomarker testing.(29–31) This top-down strategy checked for consistency with the existing literature to facilitate comparisons among populations. During a pilot/calibration phase, deductive codes were applied to the first ten open-ended responses to test their fit with the data, with codes adjusted as necessary, as raters reviewed each other’s coding and chose consensus ratings. Codes were applied to textual units of any size reflecting a consistent idea.

The codebook was treated as an unconstrained categorization matrix.(32) Deductive codes were used for data that fit this categorization; new inductive codes were created for any other data. Following calibration, raters coded the remaining 68 responses and held two consensus sessions. Between these sessions, raters updated their coded data to reflect new/adjusted codes. Inter-rater reliability was not calculated. During these sessions, raters developed and refined codes by suggesting alternate definitions, distinctions, and examples for the updated codebook. Raters also shared individual reflections on response types or trends, including claims about the normative status of biomarker disclosure (obligatory, permitted, non-obligatory). Raters synthesized these claims as a higher-level binary category to highlight respondents’ normative attitudes toward IRR return.

## 3. Results

The overall response rate for the survey was 27.1% (152/560). Though no demographic/clinical characteristics were collected, USCC participants are middle-aged (48.5 years), majority female (63.4%), predominantly white (91.3%), and highly educated (15.6 years) (Table 1).

**Table 1:** Summary of USCC participant demographics, divided by *NOTCH3* variant gene-status.

| **Demographics** | **Other**<br>**(N=101)** | **Control**<br>**(N=116)** | **Case**<br>**(N=343)** | **Total**<br>**(N=560)** |
| --- | --- | --- | --- | --- |
| **Mean age (SD)** | 47.8 (16.7) | 46.3 (15.0) | 49.4 (13.4) | 48.5 (14.4) |
| **Gender, n (%)** |  |  |  |  |
| Female | 68 (67.3%) | 76 (65.5%) | 211 (61.5%) | 355 (63.4%) |
| Male | 33 (32.7%) | 40 (34.5%) | 132 (38.5%) | 205 (36.6%) |
| **Mean years of**<br>**education (SD)** | 15.8 (2.4) | 16.0 (2.2) | 15.4 (2.4) | 15.6 (2.4) |
| **Education (Bachelor's**<br>**level), n (%)** |  |  |  |  |
| Fewer than 16 years | 38 (37.6%) | 32 (27.6%) | 154 (44.9%) | 224 (40.0%) |
| At least 16 years | 63 (62.4%) | 84 (72.4%) | 189 (55.1%) | 336 (60.0%) |
| **Race, n (%)** |  |  |  |  |
| Non-White | 9 (8.9%) | 7 (6.1%) | 32 (9.5%) | 48 (8.7%) |
| White | 92 (91.1%) | 108 (93.9%) | 306 (90.5%) | 506 (91.3%) |
| Missing | 0 | 1 | 5 | 6 |
| **Ethnicity, n (%)** |  |  |  |  |
| Not Hispanic or Latino | 95 (95.0%) | 113 (97.4%) | 307 (89.5%) | 515 (92.1%) |
| Hispanic or Latino | 5 (5.0%) | 3 (2.6%) | 36 (10.5%) | 44 (7.9%) |
| Missing | 1 | 0 | 0 | 1 |

Note: Other (*NOTCH3* variant gene-status pending); Control (*NOTCH3* variant gene-status negative); Case (*NOTCH3* variant gene-status positive).

When asked about reasons to return IRR, 97.3% of USCC participants endorsed “learn about my disease and its predicted course” as extremely/very important (mean: 4.68) (Table 2). 91.3% rated “inform lifestyle changes I might make” as extremely/very important (mean: 4.56). 87.8% endorsed “prepare my family for possible illness in the future” as extremely/very important (mean: 4.51). Respondents rated considerations of convenience (not repeating tests in different research studies [mean: 3.26] or with doctor [mean: 3.46]) less highly than these cognitive or practical considerations related to being informed or making plans.

**Table 2:** Results for survey items 1-3 (closed-ended, Likert scale 1-5), with percentages of strong endorsement and mean item values.

| **Survey Item 1: How important are these reasons to share research results?** | **Extremely or very important (%)** | **Mean (SD)** |
| --- | --- | --- |
| Learn about my disease and its predicted course | 97.3* | 4.68 (0.64) |
| Inform lifestyle changes I might make, such as diet or exercise that might help prevent declines in thinking and functioning | 91.3<sup>†</sup> | 4.56 (0.83) |
| Learn about my disease to inform my kids and family | 90.1* | 4.51 (0.87) |
| Prepare my family for possible illness in the future | 87.8<sup>†</sup> | 4.32 (1.02) |
| Participate in clinical trials attempting to improve thinking and function | 86.4<sup>†</sup> | 4.35 (0.88) |
| Put my mind at ease to know how I am doing | 83.0<sup>†</sup> | 4.33 (0.99) |
| Prepare for how I want to live with this disease | 78.2<sup>†</sup> | 4.20 (1.09) |
| Start doing things sooner than I had planned | 76.9<sup>†</sup> | 4.15 (1.11) |
| Confirm that I might already be developing symptoms of cognitive decline or dementia | 70.7<sup>†</sup> | 3.98 (1.24) |
| Arrange my personal affairs, such as insurance, my will, or finances | 64.6<sup>†</sup> | 3.85 (1.20) |
| Avoid having the same measure or test repeated by my doctor | 53.0<sup>†</sup> | 3.46 (1.36) |
| Avoid having the same measure or test repeated in different research studies | 45.9* | 3.26 (1.33) |
| **Survey Item 2: How important is it to share this type of information?** | **Extremely or very important (%)** | **Mean (SD)** |
| Imaging or MRI scans | 89.3<sup>§</sup> | 4.62 (0.76) |
| Cognitive or neuropsychological testing | 89.3<sup>§</sup> | 4.52 (0.79) |
| Neurological, motor, or sensory findings | 85.3<sup>§</sup> | 4.44 (0.88) |
| Overall diagnostic category (presymptomatic and cognitively normal; prodromal; mildly, moderately, or severely cognitively impaired) | 84.6<sup>§</sup> | 4.40 (0.84) |
| Functional capacity measures (ability to perform activities of daily living) | 84.6<sup>§</sup> | 4.40 (0.87) |
| Genetic testing results (mild or severe gene mutation) | 84.5<sup>‡</sup> | 4.45 (0.93) |
| Quality of life measures | 83.9<sup>§</sup> | 4.39 (0.96) |
| Genetic testing results (mutation or no mutation) | 83.2<sup>§</sup> | 4.34 (1.04) |
| Emotional, mood, and motivational measures | 76.5<sup>§</sup> | 4.28 (0.95) |
| Biofluid analyses (blood, saliva, cerebrospinal fluid) | 72.3<sup>‡</sup> | 4.19 (1.07) |
| **Survey Item 3: Imagine that your biomarker-based prognosis (future estimate of disease or** | **Extremely or very ____ (%)** | **Mean (SD)** |
| **symptoms) is shared with you. How would you feel in the cases below?** |  |  |
| Uncertain about my risk of developing a severe form of CADASIL | 24.0* | 2.82 (1.13) |
| Anxious or nervous about the results | 18.2‡ | 2.66 (1.07) |
| Difficult to share these results with others | 12.9† | 2.07 (1.15) |
| Concerned about other people treating me differently if they learned the results | 12.2‡ | 2.03 (1.19) |

Note: \*n=146, †n=147, ‡n=148, §n=149.

When asked about the importance of returning different data types, 89.3% of USCC participants endorsed “imaging or MRI scans” (mean: 4.62) and “cognitive/neuropsychological testing” (mean: 4.52) as extremely/very important to return (Table 2). Overall, participants rated all IRR as extremely/very important to return, with the lowest rated type (“biofluid analyses [blood/saliva/cerebrospinal fluid]) endorsed by 72.3% (mean: 4.19).

When asked to imagine their reaction in a scenario wherein their biomarker-based prognosis (future estimate of disease or symptoms) were shared, 24% of participants endorsed feeling extremely/very uncertain about their risk for severe CADASIL forms (mean: 2.82), 18.2% extremely/very anxious or nervous about results (mean: 2.66), 12.9% extremely/very difficult to share results with others (mean: 2.07), and 12.2% extremely/very concerned about being treated differently (mean: 2.03) (Table 2). Put simply, participants rated hypothetical negative outcomes as “a little” to “somewhat” concerning.

According to the Mann-Whitney U Test, USCC and ADRC/WRAP participants differed significantly in their importance ratings of eight reasons for returning IRR. Statistical testing indicates that USCC participants rated importance of reasons to learn IRR higher than ADRC/WRAP participants for seven of eight responses (*P*≤.05) (Table 3).

**Table 3:** Comparison of CADASIL and AD population responses to equivalent items from closed-ended Survey Item 1 (How important it is to receive IRR to…)

| Survey item | CADASIL<br>participant<br>rating<br>(mean) (SD)<br>(N) | AD<br>participant<br>rating<br>(mean) (SD)<br>(N) | P value |
| --- | --- | --- | --- |
| How important it is to receive IRR to... |  |  |  |
| ...learn about my disease and its predicted course | 4.68 (0.64)<br>(146) | 3.93 (1.20)<br>(331) | <.001* |
| ...inform lifestyle changes I might make, such as diet or exercise that might help prevent declines in thinking and functioning | 4.56 (0.83)<br>(147) | 4.26 (1.00)<br>(334) | <.001* |
| ...put my mind at ease to know how I am doing | 4.33 (0.99)<br>(147) | 4.13 (1.00)<br>(334) | .015* |
| ...participate in clinical trials attempting to improve thinking and function | 4.35 (0.88)<br>(147) | 3.91 (1.00)<br>(332) | <.001* |
| ...arrange my personal affairs, such as insurance, my will, or finances | 3.85 (1.20)<br>(147) | 3.73 (1.31)<br>(334) | .392 |
| ...confirm that I might already be developing symptoms of cognitive decline or dementia | 3.98 (1.24)<br>(147) | 3.77 (1.20)<br>(333) | .031* |
| ...prepare my family for possible illness in the future | 4.32 (1.02)<br>(147) | 3.99 (1.10)<br>(334) | <.001* |
| ...start doing things sooner than I had planned | 4.15 (1.11)<br>(147) | 3.77 (1.20)<br>(334) | <.001* |
| *Statistically significant |  |  |  |

Note: \*Statistically significant ( $P \leq .05$ ).

The fourth survey item invited respondents to share anything else about why study participants should or should not be able to receive their individual research findings. The response rate to the fourth item was 77/152 (50.7%). The codes with the most occurrences were “IRR return will improve my health or well-being” (23 occurrences), “IRR return will help me plan for the future” (21), and “IRR return will provide me valuable information” (17) (total: 61/135 occurrences) (Table 4). The codes with the lowest occurrences were “IRR return will make me feel good for helping others” (2), “IRR return will improve my spiritual well-being” (2), and “IRR return will raise concerns of psychological harms” (1) (5/135 occurrences). For high-level categories regarding the normative status of IRR return, 85.7% of respondents (66/77) expressed support for qualified disclosure: IRR should be returned in at least some cases to interested participants (Table 4). Respondents sometimes appealed to explicitly normative notions – informational rights, dignity, justice – to justify returning IRR.

**Table 4:** Summary of occurrences from codes and categories for survey item 4: Would you like to share anything else with the research team about why study participants should or should not be able to receive their individual research findings?

| **Codes** | **Occurrences** | **Sample Quotations** |
| --- | --- | --- |
| IRR return will improve my health or well-being | 23 | "I strongly feel that my results should be released to my CADASIL physician. These results would help her treat me and learn from my specific case." |
| IRR return will help me plan for the future | 21 | "It offers a pragmatic approach to planning for the future" |
| IRR return will provide me valuable information | 17 | "I think it is very important for the information gathered in these studies to be able to given to the participants so that they have the maximum information possible." |
| IRR return will improve my knowledge of my condition | 14 | "My neurologist is not a specialist in cadasil, but with this specialist data, we could both potentially better understand the course of the disease specifically for me as a patient." |
| IRR return will satisfy my rights to my information | 14 | "I feel like it's my body and data and i should own it or have access to it" |
| IRR return will help me cope with my situation | 11 | "It would put minds to rest knowing the facts" |
| IRR return will demonstrate justice, fairness, or reciprocity | 8 | "They [study participants] gave their time and data to help the research, and it's only fair they know what was found about them." |
| No further information provided | 8 | "I am thoroughly thankful for everyone in this study." |
| IRR return will alleviate research participant burden | 5 | "Having access to my own test results would also reduce the chance of having to duplicate different tests that aren't the most fun to endure" |
| IRR return will promote respect for persons and the research community | 4 | "It is a disservice to us as human beings to not be given the option to see our results." |
| IRR return will facilitate family communication and social support | 3 | "I wish I could help her [my sister] with important steps to control her life" |
| IRR return will raise concerns of stigma or discrimination | 3 | "[...] if I get the results then insurance might find out and thus be able to deny me for medical coverage and life insurance." |
| IRR return will make me feel good for helping others | 2 | "Thanks for [...] giving future generations options! Let's cure this!" |
| IRR return will improve my spiritual well-being | 1 | "[...] the possibility of making a positive life change." |
| IRR return will raise concerns of psychological harms | 1 | "It was more emotionally and mentally difficult to find out I was positive than I expected." |
| **Categories** |  |  |
| IRR should at least sometimes be returned to participants. | 66 | "As long as it is an option for individuals to not learn their results, my preference would be to release everything to participants that want it." |
| Insufficient information for researchers to determine attitude toward IRR return. | 8 | "No, Thank you for this opportunity!" |
| IRR need not be returned to participants. | 3 | "I feel quite torn about how much more information I want and thus might feel I had to disclose if asked." |

## 4. Discussion

This study expands the existing literature on stakeholder attitudes toward IRR by surveying a previously unstudied population: persons affected by CADASIL. Respondents rated highly many reasons for returning IRR and all data types, while rating low any concerns about their reactions to biomarker-based prognostic information. When prompted for additional information about IRR, respondents highlighted both instrumental and intrinsic value. Importantly, these results suggest that, relative to ADRC/WRAP participants, USCC respondents placed more importance on returning IRR. To our knowledge, this is the first study to directly compare different participant communities in observational neurodegenerative research.

Notwithstanding these differences, USCC and ADRC WRAP participants largely agree about IRR utility and low risks secondary to disclosure. Quantitative and qualitative results indicate that both groups rate especially highly the cognitive and practical rationales for returning IRR. In other studies, community stakeholders – participants, companions, caregivers, family – regularly endorse that disclosure promotes subjective health outcomes(24, 33–39) or opportunities for short- or long-term planning.(24, 34, 37–42) Regarding risks secondary to disclosure, the response trend is consistent with published findings. Community stakeholders reflecting on IRR typically identify more advantages and may discount potential negative consequences of disclosure.(36, 40, 41, 43) Other studies also find that attitudes to study enrollment or biomarker disclosure do not appear to vary by IRR type.(11, 44)

Despite the overall consistency with the literature, the combination of significant attitudinal differences between USCC and ADRC/WRAP participants and the dearth of research on non-genetic biomarker disclosure in dominantly inherited neurodegenerative disease supports that more research is needed about the attitudes of non-AD populations. Whereas persons affected by AD typically have a late-onset amnestic presentation with cognitive, functional, or behavioral changes, persons affected by CADASIL have earlier clinical symptoms, including cerebrovascular events, migraine with aura, cognitive decline, and behavioral changes, with differences in clinical testing and management.

In preclinical AD, amyloid-beta PET imaging and *APOE* genetic testing are probabilistic measures indicating a person’s increased lifetime risk of dementia, with clinical symptoms emerging later or even never.(45) Therapies targeting amyloid-beta are available for individuals with cognitive impairment and demonstrate small though significant clinical effects. Though *NOTCH3* testing for CADASIL is deterministic, severity staging systems and evidence-based therapies remain constrained by limited sample sizes and disease heterogeneity.(46, 47) Finally, whereas multiple organizations advocate for persons affected by AD, organizations advocating for the CADASIL community are still emerging. These CADASIL-AD differences resemble those observed among other communities, e.g., persons living with autosomal dominant FTD.(40) CADASIL community attitudes may meaningfully diverge from AD community attitudes, within certain limits. Follow-up study is therefore warranted in CADASIL and may yield new findings relevant to other adult-onset, dominantly inherited neurodegenerative diseases.

We highlight three lessons for biomarker disclosure research. First, study methods unsurprisingly influence reported outcomes. Our quantitative and qualitative results did not always align. For example, respondents endorsed “Learn about my disease to inform my kids and family” as an extremely/very important reason to share IRR (90.1% Table 2) more frequently than the code “Facilitate family communication and social support” appears in open-ended responses (3/135 occurrences, Table 4).

Second, we found strong endorsement for returning all IRR data types, including four types about which participants have never been surveyed: neurological, motor, or sensory; functional capacity; quality of life; emotional, mood, and motivational. Such strong endorsement supports the observation that this may not signal interest in specific IRR as individually meaningful informational units. Instead, it signals a desire for synthetic informational risk-packages to support stakeholders’ cognitive, practical, and affective aims.(44) Relatedly, the desired data may not clearly advance participants’ stated goals for IRR (e.g., “learn about my disease and its predicted course”, “inform lifestyle changes I might make”) more than already available information, i.e., *NOTCH3* gene-status or population-level risk for cerebrovascular events. This suggests perceived utility beyond participants’ explicitly stated goals.

Third, our findings suggest future research directions in CADASIL community engagement. A sizeable minority of USCC participants are interested in receiving IRR. To define IRR guidance for this community, a participatory process should integrate lessons from AD disclosure science and elicit CADASIL-specific considerations. Such guidance could better reflect CADASIL community stakeholder attitudes and support future trial success. Our results may also partly satisfy ethical and epistemic obligations to investigate the desirability of IRR return.(48)

Our interpretation comes with several significant limitations. Most importantly, external validity is limited by self-selection bias (27% response rate). Those participants motivated to complete the survey are likely those most invested in IRR return. We cannot rule out that statistically significant differences in the distribution of responses between the USCC and ADRC/WRAP surveys are partly derivative of respondent bias. Nevertheless, response rates are often low in similar surveys, and distributions may also be consistent with differences in respondents’ mean age.(24) Second, we did not collect any demographic/clinical characteristics for respondents. Since these characteristics may influence individual interest in IRR return, we cannot control for the their influence on responses. Like other natural history studies, the USCC is not fully representative of the US or global populations. Participants are predominantly White, financially secure, well-educated, and socialized in the US cultural context. Finally, the survey did not directly ask participants whether they want to receive IRR and may have biased responses by implying that participants want or should want IRR. These considerations limit the internal and external validity of our findings.

In sum, this study explored the attitudes of persons living with CADASIL toward IRR return through a four-item survey. USCC survey respondents expressed strong support for returning IRR, discounted the risk of negative consequences, and underscored the utility of IRR. Future study is necessary to determine whether support for IRR return is consistently high in CADASIL populations and higher than other populations. Nonetheless, this study expands our understanding of the personal utility of IRR and the informational needs of participants affected by adult-onset, dominantly inherited neurodegenerative diseases.

## Data Availability

Data produced in the present study may be made available upon reasonable request to the authors.

## Acknowledgments

We acknowledge and appreciate the valuable contributions of USCC participants, families, and research staff to advance the state of CADASIL research: https://cadasil.wisc.edu/wp-content/uploads/sites/2133/2026/08/CADASIL_Consortium_Acknowledgments.pdf. We thank study staff Jenna Spring, Ivan Griego, and Megan Caruso for assistance with advertising the survey to study participants.

## Consent statement

All participants in the USCC provided written informed consent to engage in the research study.

## Author contributions

Conceptualization: DKB, JSP

Methodology: DKB, JSP

Formal analysis: DKB, EP

Writing - Original Draft: DKB

Writing - Review & Editing: DKB, EP, LRC, FBK, KDC, JSP

Supervision: JSP

Project administration: DKB

Funding acquisition: JSP

## References

1. Bookman EB, Langehorne AA, Eckfeldt JH, Glass KC, Jarvik GP, Klag M, et al. Reporting genetic results in research studies: summary and recommendations of an NHLBI working group. Am J Med Genet A. 2006;140(10):1033–40.

2. National Academies of Sciences E, and Medicine,. Returning Individual Research Results to Participants: Guidance for a New Research Paradigm. Washington (DC): National Academies of Sciences, Engineering, and Medicine,; 2018. Report No.: 978-0-309-47517-4.

3. National Bioethics Advisory Commission. Research Involving Human Biological Materials: Ethical Issues and Policy Guidance Volume 1: Report and Recommendations of the NBAC. Rockville, MD; 1999.

4. National Heart Lung Blood Institute Working Group, Fabsitz RR, McGuire A, Sharp RR, Puggal M, Beskow LM, et al. Ethical and practical guidelines for reporting genetic research results to study participants: updated guidelines from a National Heart, Lung, and Blood Institute working group. Circ Cardiovasc Genet. 2010;3(6):574–80.

5. Presidential Commission for the Study of Bioethical Issues. Anticipate and communicate: ethical management of incidental and secondary findings in the clinical, research, and direct-to-consumer contexts. Washington, District of Columbia: Presidential Commission for the Study of Bioethical Issues,; 2013. Available from: HeinOnline Reports of U.S. Presidential Commissions https://www.heinonline.org/HOL/Page?handle=hein.prescomm/prescommaaabk0001&id=1&size=2&collection=prescomm&index=prescommcom.

6. Secretary’s Advisory Committee on Human Research Protections. Letter to Secretary of Health and Human Services, the Honorable Sylvia Burwell. Attachment C: Return of Individual Results and Special Consideration of Issues Arising from Amendments of HIPAA and CLIA: Secretary’s Advisory Committee on Human Research Protections,; 2015 [Available from: http://www.hhs.gov/ohrp/sachrp-committee/recommendations/2015-september-28-attachment-c/index.html.

7. The Multi-Regional Clinical Trials (MRCT) Center. Return of Individual Results to Participants: Recommendations Document. Cambridge, MA: The Multi-Regional Clinical Trials (MRCT) Center,; 2017. Available from: https://mrctcenter.org/resource/return-of-individual-results-to-participants-recommendations-document-version-1-2/.

8. Walter S, Taylor A, Tyrone J, Langer S, Pagan JR, Hummel CH, et al. Disclosing Individual Results in Dementia Research: A Proposed Study Participant’s Bill of Rights. J Alzheimers Dis. 2022;90(3):945–52.

9. FDA CDER and CBER Working Group. Qualification Process for Drug Development Tools: Guidance for Industry and FDA Staff Silver Spring (MD) Bethesda (MD): DHHS; 2020 [Available from: https://www.fda.gov/regulatory-information/search-fda-guidance-documents/qualification-process-drug-development-tools-guidance-industry-and-fda-staff.

10. van der Schaar J, Visser LNC, Asscher ECA, Pijnenburg YAL, de Geus CM, van der Flier WM, et al. Impact of diagnostic genetic testing for familial dementia: experiences of patients and relatives. Alzheimers Res Ther. 2026;18(1).

11. Eliacin J, Hathaway E, Wang S, O’Connor C, Saykin AJ, Cameron KA. Factors influencing the participation of Black and White Americans in Alzheimer’s disease biomarker research. Alzheimers Dement (Amst). 2022;14(1):e12384.

12. Passmore SR, Longhurst C, Gerbitz A, Green-Harris G, Norris N, Edwards DF. “I Want to Know Everything … “: The Return of Research Results and the Importance of Transparency in the Acceptability of Lumbar Punctures for African American Older Adults. J Alzheimers Dis. 2023;95(2):663–75.

13. Zhou Y, Elashoff D, Kremen S, Teng E, Karlawish J, Grill JD. African Americans are less likely to enroll in preclinical Alzheimer’s disease clinical trials. Alzheimers Dement (N Y). 2017;3(1):57–64.

14. Rahman-Filipiak A, Chin NA, Kohl H, Reader JM, Erickson CM, Dickerson BC, et al. Return of research results across the Alzheimer’s Disease Research Centers network. Alzheimers Dement. 2025;21(6):e70418.

15. Bollinger RM, Gabel M, Coble DW, Chen SW, Keleman AA, Doralus J, et al. Retention of Study Partners in Longitudinal Studies of Alzheimer Disease. J Alzheimers Dis. 2023;94(1):189–99.

16. Erickson CM, Chin NA, Johnson SC, Gleason CE, Clark LR. Disclosure of preclinical Alzheimer’s disease biomarker results in research and clinical settings: Why, how, and what we still need to know. Alzheimers Dement (Amst). 2021;13(1):e12150.

17. Gabel M, Bollinger RM, Coble DW, Grill JD, Edwards DF, Lingler JH, et al. Retaining Participants in Longitudinal Studies of Alzheimer’s Disease. J Alzheimers Dis. 2022;87(2):945–55.

18. Grill JD, Zhou Y, Elashoff D, Karlawish J. Disclosure of amyloid status is not a barrier to recruitment in preclinical Alzheimer’s disease clinical trials. Neurobiol Aging. 2016;39:147–53.

19. Ketchum FB, Erickson CM, Chin NA, Gleason CE, Lambrou NH, Benton SF, et al. What Influences the Willingness of Blacks and African Americans to Enroll in Preclinical Alzheimer’s Disease Biomarker Research? A Qualitative Vignette Analysis. J Alzheimers Dis. 2022;87(3):1167–79.

20. Kolarcik CL, Bledsoe MJ, O’Leary TJ. Returning Individual Research Results to Vulnerable Individuals. Am J Pathol. 2022;192(9):1218–29.

21. Molinuevo JL, Cami J, Carne X, Carrillo MC, Georges J, Isaac MB, et al. Ethical challenges in preclinical Alzheimer’s disease observational studies and trials: Results of the Barcelona summit. Alzheimers Dement. 2016;12(5):614–22.

22. Chabriat H, Joutel A, Dichgans M, Tournier-Lasserve E, Bousser MG. Cadasil. Lancet Neurol. 2009;8(7):643–53.

23. Hack RJ, Gravesteijn G, Cerfontaine MN, Santcroos MA, Gatti L, Kopczak A, et al. Three-tiered EGFr domain risk stratification for individualized NOTCH3-small vessel disease prediction. Brain. 2023;146(7):2913–27.

24. Clark LR, Erickson CM, Jonaitis EM, Ma Y, Chin NA, Basche K, et al. Anticipated reactions to learning Alzheimer’s disease biomarker results. Alzheimers Res Ther. 2022;14(1):85.

25. Roberts JS, LaRusse SA, Katzen H, Whitehouse PJ, Barber M, Post SG, et al. Reasons for seeking genetic susceptibility testing among first-degree relatives of people with Alzheimer disease. Alzheimer Dis Assoc Disord. 2003;17(2):86–93.

26. Roberts JS, Ferber R, Blacker D, Rumbaugh M, Grill JD, Advisory Group on Risk Evidence Education for D. Disclosure of individual research results at federally funded Alzheimer’s Disease Research Centers. Alzheimers Dement (N Y). 2021;7(1):e12213.

27. Fereday J, Muir-Cochrane E. Demonstrating Rigor Using Thematic Analysis: A Hybrid Approach of Inductive and Deductive Coding and Theme Development. International Journal of Qualitative Methods. 2006;5(1):80–92.

28. Crabtree BF, Miller WL. Doing Qualitative Research: SAGE Publications; 1999.

29. Kohler JN, Turbitt E, Biesecker BB. Personal utility in genomic testing: a systematic literature review. Eur J Hum Genet. 2017;25(6):662–8.

30. Kohler JN, Turbitt E, Lewis KL, Wilfond BS, Jamal L, Peay HL, et al. Defining personal utility in genomics: A Delphi study. Clin Genet. 2017;92(3):290–7.

31. Smedinga M, Tromp K, Schermer MHN, Richard E. Ethical Arguments Concerning the Use of Alzheimer’s Disease Biomarkers in Individuals with No or Mild Cognitive Impairment: A Systematic Review and Framework for Discussion. J Alzheimers Dis. 2018;66(4):1309–22.

32. Elo S, Kyngas H. The qualitative content analysis process. J Adv Nurs. 2008;62(1):107–15.

33. Hendriksen HMA, de Rijke TJ, Fruijtier A, van de Giessen E, van Harten AC, van Leeuwenstijn-Koopman M, et al. Amyloid PET disclosure in subjective cognitive decline: Patient experiences over time. Alzheimers Dement. 2024;20(9):6556–65.

34. Lohmeyer JL, Alpinar-Sencan Z, Schicktanz S. Attitudes towards prediction and early diagnosis of late-onset dementia: a comparison of tested persons and family caregivers. Aging Ment Health. 2021;25(5):832–43.

35. Milne R, Bunnik E, Diaz A, Richard E, Badger S, Gove D, et al. Perspectives on Communicating Biomarker-Based Assessments of Alzheimer’s Disease to Cognitively Healthy Individuals. J Alzheimers Dis. 2018;62(2):487–98.

36. Rahman-Filipiak A, Lesniak M, Burgei A, Sadaghiyani S, Roberts JS, Lichtenberg PA, et al. Qualitative insights into participant and care partner perspectives on research-based Alzheimer’s disease biomarker testing and disclosure. J Alzheimers Dis Rep. 2026;10:25424823261429997.

37. Rahman-Filipiak A, Lesniak M, Sadaghiyani S, Roberts S, Lichtenberg P, Hampstead BM. Perspectives From Black and White Participants and Care Partners on Return of Amyloid and Tau PET Imaging and Other Research Results. Alzheimer Dis Assoc Disord. 2023;37(4):274–81.

38. Smith HS, Robinson JO, Levchenko A, Pereira S, Pascual B, Bradbury K, et al. Research Participants’ Perspectives on Precision Diagnostics for Alzheimer’s Disease. J Alzheimers Dis. 2024;97(3):1261–74.

39. Vanderschaeghe G, Schaeverbeke J, Vandenberghe R, Dierickx K. Amnestic MCI Patients’ Perspectives toward Disclosure of Amyloid PET Results in a Research Context. Neuroethics. 2017;10(2):281–97.

40. Graafland CH, Seelaar H, Panman JL, Jiskoot LC, Kleefstra T, Poos JM, et al. “It seems enormously valuable to me.” Perspectives of Dutch (potential) carriers of genetic FTD on onset-predictive biomarker testing. Alzheimers Res Ther. 2025;17(1):99.

41. Lingler JH, Roberts JS, Kim H, Morris JL, Hu L, Mattos M, et al. Amyloid positron emission tomography candidates may focus more on benefits than risks of results disclosure. Alzheimers Dement (Amst). 2018;10:413–20.

42. Vanderschaeghe G, Vandenberghe R, Dierickx K. Stakeholders’ Views on Early Diagnosis for Alzheimer’s Disease, Clinical Trial Participation and Amyloid PET Disclosure: A Focus Group Study. J Bioeth Inq. 2019;16(1):45–59.

43. Vanderschaeghe G, Schaeverbeke J, Bruffaerts R, Vandenberghe R, Dierickx K. Amnestic MCI patients’ experiences after disclosure of their amyloid PET result in a research context. Alzheimers Res Ther. 2017;9(1):92.

44. Mozersky J, Hartz S, Linnenbringer E, Levin L, Streitz M, Stock K, et al. Communicating 5-Year Risk of Alzheimer’s Disease Dementia: Development and Evaluation of Materials that Incorporate Multiple Genetic and Biomarker Research Results. J Alzheimers Dis. 2021;79(2):559–72.

45. Rabinovici GD, Knopman DS, Arbizu J, Benzinger TLS, Donohoe KJ, Hansson O, et al. Updated appropriate use criteria for amyloid and tau PET: A report from the Alzheimer’s Association and Society for Nuclear Medicine and Molecular Imaging Workgroup. Alzheimers Dement. 2025;21(1):e14338.

46. Gravesteijn G, Rutten JW, Cerfontaine MN, Hack RJ, Liao YC, Jolly AA, et al. Disease Severity Staging System for NOTCH3-Associated Small Vessel Disease, Including CADASIL. JAMA Neurol. 2025;82(1):49–60.

47. Meschia JF, Worrall BB, Elahi FM, Ross OA, Wang MM, Goldstein ED, et al. Management of Inherited CNS Small Vessel Diseases: The CADASIL Example: A Scientific Statement From the American Heart Association. Stroke. 2023;54(10):e452–e64.

48. Burks DK, Paulsen JS. Ethical challenges to the responsible communication of individual results from observational neurodegeneration research. Developments in Neuroethics and Bioethics: Academic Press; 2026.

